# Genetic architectures of proximal and distal colorectal cancer are partly distinct

**DOI:** 10.1101/2020.05.01.20087957

**Authors:** Jeroen R Huyghe, Tabitha A Harrison, Stephanie A Bien, Heather Hampel, Jane C Figueiredo, Stephanie L Schmit, David V Conti, Sai Chen, Conghui Qu, Yi Lin, Richard Barfield, John A Baron, Amanda J Cross, Brenda Diergaarde, David Duggan, Sophia Harlid, Liher Imaz, Hyun Min Kang, David M Levine, Vittorio Perduca, Aurora Perez-Cornago, Lori C Sakoda, Fredrick R Schumacher, Martha L Slattery, Amanda E Toland, Franzel JB van Duijnhoven, Bethany Van Guelpen, Volker Arndt, Antonio Agudo, Demetrius Albanes, M Henar Alonso, Kristin Anderson, Coral Arnau-Collell, Barbara Banbury, Michael C Bassik, Sonja I Berndt, Stéphane Bézieau, D Timothy Bishop, Juergen Boehm, Heiner Boeing, Marie-Christine Boutron-Ruault, Hermann Brenner, Stefanie Brezina, Stephan Buch, Daniel D Buchanan, Andrea Burnett-Hartman, Bette J Caan, Peter T Campbell, Prudence Carr, Antoni Castells, Sergi Castellví-Bel, Andrew T Chan, Jenny Chang-Claude, Stephen J Chanock, Keith R Curtis, Albert de la Chapelle, Douglas F Easton, Dallas R English, Edith JM Feskens, Manish Gala, Steven J Gallinger, W James Gauderman, Graham G Giles, Phyllis J Goodman, William M Grady, John S Grove, Andrea Gsur, Marc J Gunter, Robert W Haile, Jochen Hampe, Michael Hoffmeister, John L Hopper, Wan-Ling Hsu, Wen-Yi Huang, Thomas J Hudson, Mazda Jenab, Mark A Jenkins, Amit D Joshi, Temitope O Keku, Charles Kooperberg, Tilman Kuhn, Sébastien Küry, Loic Le Marchand, Flavio Lejbkowicz, Christopher I Li, Li Li, Wolfgang Lieb, Annika Lindblom, Noralane M Lindor, Satu Männistö, Sanford D Markowitz, Roger L Milne, Lorena Moreno, Neil Murphy, Rami Nassir, Kenneth Offit, Shuji Ogino, Salvatore Panico, Patrick S Parfrey, Rachel Pearlman, Paul D P Pharoah, Amanda I Phipps, Elizabeth A Platz, John D Potter, Ross L Prentice, Lihong Qi, Leon Raskin, Gad Rennert, Hedy S Rennert, Elio Riboli, Clemens Schafmayer, Robert E Schoen, Daniela Seminara, Mingyang Song, Yu-Ru Su, Catherine M Tangen, Stephen N Thibodeau, Duncan C Thomas, Antonia Trichopoulou, Cornelia M Ulrich, Kala Visvanathan, Pavel Vodicka, Ludmila Vodickova, Veronika Vymetalkova, Korbinian Weigl, Stephanie J Weinstein, Emily White, Alicja Wolk, Michael O Woods, Anna H Wu, Goncalo R Abecasis, Deborah A Nickerson, Peter C Scacheri, Anshul Kundaje, Graham Casey, Stephen B Gruber, Li Hsu, Victor Moreno, Richard B Hayes, Polly A Newcomb, Ulrike Peters

**Affiliations:** Public Health Sciences Division, Fred Hutchinson Cancer Research Center, Seattle, Washington, USA; Division of Human Genetics, Department of Internal Medicine, The Ohio State University Comprehensive Cancer Center, Columbus, Ohio, USA; Department of Medicine, Samuel Oschin Comprehensive Cancer Institute, Cedars-Sinai Medical Center, Los Angeles, California, USA; Department of Preventive Medicine, Keck School of Medicine, University of Southern California Los Angeles, California, USA; Department of Cancer Epidemiology, H. Lee Moffitt Cancer Center and Research Institute, Tampa, Florida, USA; Department of Preventive Medicine, USC Norris Comprehensive Cancer Center, Keck School of Medicine, University of Southern California, Los Angeles, California, USA; Department of Biostatistics and Center for Statistical Genetics, University of Michigan, Ann Arbor, Michigan, USA; Department of Medicine, University of North Carolina School of Medicine, Chapel Hill, North Carolina, USA; Department of Epidemiology and Biostatistics, Imperial College London, London, UK; Department of Human Genetics, Graduate School of Public Health, University of Pittsburgh, and UPMC Hillman Cancer Center, Pittsburgh, PA; Translational Genomics Research Institute - An Affiliate of City of Hope, Phoenix, Arizona, USA; Department of Radiation Sciences, Oncology Unit, Umeå University, Umeå, Sweden; Public Health Division of Gipuzkoa, Health Department of Basque Country, Spain; Department of Biostatistics, University of Washington, Seattle, Washington, USA; Laboratoire de Mathématiques Appliquées MAP5 (UMR CNRS 8145), Université Paris Descartes, Paris, France; CESP (Inserm U1018), Facultés de Médicine Université Paris-Sud, UVSQ, Université Paris-Saclay, Gustave Roussy, 94805, Villejuif, France; Cancer Epidemiology Unit, Nuffield Department of Population Health, University of Oxford, Oxford, UK; Division of Research, Kaiser Permanente Northern California, Oakland, California, USA; Department of Population and Quantitative Health Sciences, Case Western Reserve University, Cleveland, Ohio, USA; Department of Internal Medicine, University of Utah, Salt Lake City, Utah, USA; Departments of Cancer Biology and Genetics and Internal Medicine, Comprehensive Cancer Center, The Ohio State University, Columbus, Ohio, USA; Division of Human Nutrition and Health, Wageningen University & Research, Wageningen, The Netherlands; Division of Clinical Epidemiology and Aging Research, German Cancer Research Center (DKFZ), Heidelberg, Germany; Unit of Nutrition and Cancer, Cancer Epidemiology Research Program, Catalan Institute of Oncology-IDIBELL, L’Hospitalet de Llobregat, Barcelona, Spain; Division of Cancer Epidemiology and Genetics, National Cancer Institute, National Institutes of Health, Bethesda, Maryland, USA; Cancer Prevention and Control Program, Catalan Institute of Oncology-IDIBELL, L’Hospitalet de Llobregat, Barcelona, Spain; CIBER de Epidemiología y Salud Pública (CIBERESP), Madrid, Spain; Department of Clinical Sciences, Faculty of Medicine, University of Barcelona, Barcelona, Spain; Division of Epidemiology and Community Health, University of Minnesota, Minneapolis, Minnesota, USA; Gastroenterology Department, Hospital Clínic, Institut d’Investigacions Biomèdiques August Pi i Sunyer (IDIBAPS), Centro de Investigación Biomédica en Red de Enfermedades Hepáticas y Digestivas (CIBEREHD), University of Barcelona, Barcelona, Spain; Department of Genetics, Stanford University, Stanford, California, USA; Service de Génétique Médicale, Centre Hospitalier Universitaire (CHU) Nantes, Nantes, France; Leeds Institute of Medical Research at St James’s, University of Leeds, Leeds, UK; Huntsman Cancer Institute and Department of Population Health Sciences, University of Utah, Salt Lake City, Utah, USA; Department of Epidemiology, German Institute of Human Nutrition (DIfE), Potsdam-Rehbrücke, Germany; Inserm U1018, Center for Research in Epidemiology and Population Health (CESP), Gustave Roussy, Villejuif, France; Paris-South Saclay University, Villejuif, France; Division of Preventive Oncology, German Cancer Research Center (DKFZ) and National Center for Tumor Diseases (NCT), Heidelberg, Germany; German Cancer Consortium (DKTK), German Cancer Research Center (DKFZ), Heidelberg, Germany; Institute of Cancer Research, Department of Medicine I, Medical University of Vienna, Vienna, Austria; Department of Medicine I, University Hospital Dresden, Technische Universität Dresden (TU Dresden), Dresden, Germany; Centre for Epidemiology and Biostatistics, Melbourne School of Population and Global Health, The University of Melbourne, Melbourne, Victoria, Australia; Colorectal Oncogenomics Group, Department of Clinical Pathology, The University of Melbourne, Parkville, Victoria, Australia; Genomic Medicine and Family Cancer Clinic, Royal Melbourne Hospital, Parkville, Victoria, Australia; Institute for Health Research, Kaiser Permanente Colorado, Denver, Colorado, USA; Division of Research, Kaiser Permanente Medical Care Program, Oakland, California, USA; Behavioral and Epidemiology Research Group, American Cancer Society, Atlanta, Georgia, USA; Division of Clinical Epidemiology, German Cancer Research Center (DKFZ), Heidelberg, Germany; Division of Gastroenterology, Massachusetts General Hospital and Harvard Medical School, Boston, Massachusetts, USA; Channing Division of Network Medicine, Brigham and Women’s Hospital and Harvard Medical School, Boston, Massachusetts, USA; Clinical and Translational Epidemiology Unit, Massachusetts General Hospital and Harvard Medical School, Boston, Massachusetts, USA; Broad Institute of Harvard and MIT, Cambridge, Massachusetts, USA; Department of Epidemiology, Harvard T.H. Chan School of Public Health, Harvard University, Boston, Massachusetts, USA; Department of Immunology and Infectious Diseases, Harvard T.H. Chan School of Public Health, Harvard University, Boston, Massachusetts, USA; Division of Cancer Epidemiology, German Cancer Research Center (DKFZ), Heidelberg, Germany; Cancer Epidemiology Group, University Medical Centre Hamburg-Eppendorf, University Cancer Centre Hamburg (UCCH), Hamburg, Germany; Department of Cancer Biology and Genetics and the Comprehensive Cancer Center, The Ohio State University, Columbus, Ohio, USA; Department of Public Health and Primary Care, University of Cambridge, Cambridge, UK; Cancer Epidemiology Division, Cancer Council Victoria, Melbourne, Victoria, Australia; Lunenfeld Tanenbaum Research Institute, Mount Sinai Hospital, University of Toronto, Toronto, Ontario, Canada; SWOG Statistical Center, Fred Hutchinson Cancer Research Center, Seattle, Washington, USA; Clinical Research Division, Fred Hutchinson Cancer Research Center, Seattle, Washington, USA; University of Hawaii Cancer Research Center, Honolulu, Hawaii, USA; Nutrition and Metabolism Section, International Agency for Research on Cancer, World Health Organization, Lyon, France; Department of Epidemiology, School of Public Health and Institute of Health and Environment, Seoul National University, Seoul, South Korea; Ontario Institute for Cancer Research, Toronto, Ontario, Canada; International Agency for Research on Cancer, World Health Organization, Lyon, France; Center for Gastrointestinal Biology and Disease, University of North Carolina, Chapel Hill, North Carolina, USA; The Clalit Health Services, Personalized Genomic Service, Carmel, Haifa, Israel; Department of Community Medicine and Epidemiology, Lady Davis Carmel Medical Center, Haifa, Israel; Clalit National Cancer Control Center, Haifa, Israel; Department of Family Medicine, University of Virginia, Charlottesville, Virginia, USA; Institute of Epidemiology, PopGen Biobank, Christian-Albrechts-University Kiel, Kiel, Germany; Department of Clinical Genetics, Karolinska University Hospital, Stockholm, Sweden; Department of Molecular Medicine and Surgery, Karolinska Institutet, Stockholm, Sweden; Department of Health Science Research, Mayo Clinic, Scottsdale, Arizona, USA; Department of Public Health Solutions, National Institute for Health and Welfare, Helsinki, Finland; Departments of Medicine and Genetics, Case Comprehensive Cancer Center, Case Western Reserve University, and University Hospitals of Cleveland, Cleveland, Ohio, USA; Department of Pathology, School of Medicine, Umm Al-Qura’a University, Saudi Arabia; Clinical Genetics Service, Department of Medicine, Memorial Sloan Kettering Cancer Center, New York, New York, USA; Department of Medicine, Weill Cornell Medical College, New York, New York, USA; Program in MPE Molecular Pathological Epidemiology, Department of Pathology, Brigham and Women’s Hospital, Harvard Medical School, Boston, Massachusetts, USA; Department of Oncologic Pathology, Dana-Farber Cancer Institute, Boston, Massachusetts, USA; Dipartimento di Medicina Clinica e Chirurgia, Federico II University, Naples, Italy; Clinical Epidemiology Unit, Faculty of Medicine, Memorial University, St. John’s, Newfoundland, Canada; Department of Epidemiology, University of Washington, Seattle, Washington, USA; Department of Epidemiology, Johns Hopkins Bloomberg School of Public Health, Johns Hopkins University, Baltimore, Maryland, USA; Department of Public Health Sciences, School of Medicine, University of California Davis, Davis, California, USA; Division of Epidemiology, Vanderbilt Epidemiology Center, Vanderbilt University School of Medicine, Nashville, Tennessee, USA; Ruth and Bruce Rappaport Faculty of Medicine, Technion-Israel Institute of Technology, Haifa, Israel; School of Public Health, Imperial College London, London, UK; Department of General Surgery, University Hospital Rostock, Rostock, Germany; Department of Medicine and Epidemiology, University of Pittsburgh Medical Center, Pittsburgh, Pennsylvania, USA; Division of Cancer Control and Population Sciences, National Cancer Institute, Bethesda, Maryland, USA; Department of Nutrition, Harvard T.H. Chan School of Public Health, Harvard University, Boston, Massachusetts, USA; Division of Laboratory Genetics, Department of Laboratory Medicine and Pathology, Mayo Clinic, Rochester, Minnesota, USA; Hellenic Health Foundation, Athens, Greece; WHO Collaborating Center for Nutrition and Health, Unit of Nutritional Epidemiology and Nutrition in Public Health, Department of Hygiene, Epidemiology and Medical Statistics, School of Medicine, National and Kapodistrian University of Athens, Greece; Department of Molecular Biology of Cancer, Institute of Experimental Medicine of the Czech Academy of Sciences, Prague, Czech Republic; Institute of Biology and Medical Genetics, First Faculty of Medicine, Charles University, Prague, Czech Republic; Faculty of Medicine and Biomedical Center in Pilsen, Charles University, Pilsen, Czech Republic; Medical Faculty, University of Heidelberg, Germany; Institute of Environmental Medicine, Karolinska Institutet, Stockholm, Sweden; Memorial University of Newfoundland, Discipline of Genetics, St. John’s, Canada; Department of Genome Sciences, University of Washington, Seattle, Washington, USA; Department of Genetics and Genome Sciences, Case Western Reserve University School of Medicine, Case Comprehensive Cancer Center, Cleveland, Ohio, USA; Department of Computer Science, Stanford University, Stanford, California, USA; Center for Public Health Genomics, University of Virginia, Charlottesville, Virginia, USA; Division of Epidemiology, Department of Population Health, New York University School of Medicine, New York, New York, USA

**Keywords:** colorectal cancer, genetic risk factors, genetic heterogeneity, proximal colon, distal colorectum

## Abstract

**Objective:** An understanding of the etiologic heterogeneity of colorectal cancer (CRC) is critical for improving precision prevention, including individualized screening recommendations and the discovery of novel drug targets and repurposable drug candidates for chemoprevention. Known differences in molecular characteristics and environmental risk factors among tumors arising in different locations of the colorectum suggest partly distinct mechanisms of carcinogenesis. The extent to which the contribution of inherited genetic risk factors for sporadic CRC differs by anatomical subsite of the primary tumor has not been examined.

**Design:** To identify new anatomical subsite-specific risk loci, we performed genome-wide association study (GWAS) meta-analyses including data of 48,214 CRC cases and 64,159 controls of European ancestry. We characterized effect heterogeneity at CRC risk loci using multinomial modeling.

**Results:** We identified 13 loci that reached genome-wide significance *(P*<5×10^−8^) and that were not reported by previous GWAS for overall CRC risk. Multiple lines of evidence support candidate genes at several of these loci. We detected substantial heterogeneity between anatomical subsites. Just over half (61) of 109 known and new risk variants showed no evidence for heterogeneity. In contrast, 22 variants showed association with distal CRC (including rectal cancer), but no evidence for association or an attenuated association with proximal CRC. For two loci, there was strong evidence for effects confined to proximal colon cancer.

**Conclusion:** Genetic architectures of proximal and distal CRC are partly distinct. Studies of risk factors and mechanisms of carcinogenesis, and precision prevention strategies should take into consideration the anatomical subsite of the tumor.

**Significance of this study:**

What is already known about this subject?
- Heterogeneity among colorectal cancer (CRC) tumors originating at different locations of the colorectum has been revealed in somatic genomes, epigenomes, and transcriptomes, and in some established environmental risk factors for CRC.
- Genome-wide association studies (GWAS) have identified over 100 genetic variants for overall CRC risk; however, a comprehensive analysis of the extent to which genetic risk factors differ by the anatomical sublocation of the primary tumor is lacking.

What are the new findings?
- In this large consortium-based study, we analyzed clinical and genome-wide genotype data of 112,373 CRC cases and controls of European ancestry to comprehensively examine whether CRC case subgroups defined by anatomical sublocation have distinct germline genetic etiologies.
- We discovered 13 new loci at genome-wide significance (*P*<5×10^−8^) that were specific to certain anatomical sublocations and that were not reported by previous GWAS for overall CRC risk; multiple lines of evidence support strong candidate target genes at several of these loci, including *PTGER3, LCT, MLH1, CDX1, KLF14, PYGL, BCL11B*, and *BMP7*.
- Systematic heterogeneity analysis of genetic risk variants for CRC identified thus far, revealed that the genetic architectures of proximal and distal CRC are partly distinct.
- Taken together, our results further support the idea that tumors arising in different anatomical sublocations of the colorectum may have distinct etiologies.

How might it impact on clinical practice in the foreseeable future?
- Our results provide an informative resource for understanding the differential role that genes and pathways may play in the mechanisms of proximal and distal CRC carcinogenesis.
- The new insights into the etiologies of proximal and distal CRC may inform the development of new precision prevention strategies, including individualized screening recommendations and the discovery of novel drug targets and repurposable drug candidates for chemoprevention.
- Our findings suggest that future studies of etiological risk factors for CRC and molecular mechanisms of carcinogenesis should take into consideration the anatomical sublocation of the colorectal tumor.

## INTRODUCTION

Despite improvements in prevention, screening and therapy, colorectal cancer (CRC) remains one of the leading causes of cancer-related death worldwide, with an estimated 53,200 fatal cases in 2020 in the United States alone.[1] CRCs that arise proximal (right) or distal (left) to the splenic flexure differ in age- and sex-specific incidence rates, clinical, pathological and tumor molecular features.[2–5] These observed differences reflect a complex interplay between differential exposure of colorectal crypt cells to local environmental carcinogenic and protective factors in the luminal content (including the microbiome), and distinct inherent biological characteristics that may influence neoplasia risk, including sex and differences between anatomical segments in embryonic origin, development, physiology, function, and mucosal immunology. The precise extrinsic and intrinsic etiologic factors involved, their relative contributions, and how they interact to influence the carcinogenic process remain largely elusive.

An individual’s genetic background plays an important role in the initiation and development of sporadic CRC. Based on twin registries, heritability is estimated to be around 35%.[6] Since genome-wide association studies (GWASs) became possible just over a decade ago, over 100 independent genetic association signals for overall sporadic CRC risk have been identified, over half of which were only identified in the past few years.[7–10] Three decades ago, based on observed similarities between Lynch syndrome and proximal sporadic CRC, and between Familial Adenomatous Polyposis (FAP) and distal sporadic CRC, Bufill proposed the existence of two distinct genetic categories of sporadic CRC according to the location of the primary tumor. [2] However, given that genetic variants that influence sporadic CRC risk typically have small effect sizes, until very recently, sample sizes of GWASs did not provide adequate statistical power to conduct meaningful subsite analyses. As a consequence, discovery GWASs to detect genetic variants that are specific for CRC case subgroups defined by anatomic subsite of the primary tumor have not been reported yet. Similarly, a comprehensive analysis of the extent to which allelic risk of the known GWAS-identified genetic variants differs by the anatomic subsite of the primary tumor is lacking.

To address the major gap in our knowledge of the differential role that genetic variants, genes and pathways play in the mechanisms of proximal and distal CRC carcinogenesis, we conducted a large consortium-based study that included clinical and genome-wide genotype data for 112,373 CRC cases and controls. First, to discover new loci and genetic risk variants with site-specific allelic effects, we conducted GWASs of CRC case subgroups defined by the location of their primary tumor within the colorectum. Next, we systematically characterized heterogeneity of allelic effects between primary tumor subsites for new and previously identified CRC risk variants to identify loci with shared and site-specific allelic effects.

## METHODS

Detailed methods are provided in the online supplementary materials.

### Samples and genotypes

This study included clinical and genotype data for 48,214 CRC cases and 64,159 controls from three consortia: the Genetics and Epidemiology of Colorectal Cancer Consortium (GECCO), the Colorectal Cancer Transdisciplinary Study (CORECT), and Colorectal Cancer Family Registry (CCFR). Supplementary table 1 provides details on sample numbers and demographic characteristics by study. All analyses were restricted to genetically inferred European-ancestry participants. Across studies, participant recruitment occurred between the early 1990s and the 2010s. Details of all genotype data sets, genotype QC, sample selection, and studies included in this analysis have been published previously.[7,8,11,12] All participants provided written informed consent, and each study was approved by the relevant research ethics committee or institutional review board.

### Colorectal tumor anatomic sublocation definitions

We defined proximal colon cancer as any primary tumor arising in the cecum, ascending colon, hepatic flexure, or transverse colon; distal colon cancer as any primary tumor arising in the splenic flexure, descending colon, or sigmoid colon; and rectal cancer as any primary tumor arising in the rectum or rectosigmoid junction. For the GWAS discovery analyses, we analyzed five case subgroups based on primary tumor sublocation. In addition to the three aforementioned mutually exclusive case sets (proximal colon, distal colon, and rectal cancer), we defined colon cancer and distal/left-sided colorectal cancer case sets. Colon cancer cases comprised combined proximal colon and distal colon cancer cases, and additional colon cases with unspecified site. In the distal/left-sided colorectal cancer cases analysis, we combined distal colon and rectal cancer cases based on the different embryonic origins of the proximal colon versus the distal colon and rectum. Supplementary figure 1 and supplementary table 1 summarize distributions of age of diagnosis by sex and primary tumor site.

### Statistical analysis

#### GWAS meta-analyses

We imputed all genotype data sets to the Haplotype Reference Consortium (HRC) panel, which by combining sequencing data from 32,488 individuals, enables accurate imputation of single nucleotide variants (SNVs) with minor allele frequencies (MAF) as low as 0.1%.[13] In brief, we phased all genotyping array data sets using SHAPEIT2[14] and used the Michigan Imputation Server[15] to impute to the HRC panel. Within each data set, variants with an imputation accuracy *r*^2^ ≥0.3 and minor allele count (MAC) ≥50 were tested for association with CRC case subgroup. Variants that only passed filters in a single genotype data set were excluded. We assumed an additive genetic model using the imputed genotype dosage in a logistic regression model adjusted for age, sex, and study or genotyping project-specific covariates, including principal components to adjust for population structure. Details of the covariate corrections for each data set have been published previously.[8] Because Wald tests can be anti-conservative for rare variants, we performed likelihood ratio tests and combined association summary statistics across sample sets via fixed-effects meta-analysis employing Stouffer’s method, implemented in the METAL software.[16] Reported *P*-values are based on this analysis. Reported combined odds ratio (OR) estimates and 95% confidence intervals (CIs) are based on an inverse variance-weighted fixed-effects meta-analysis.

#### Heterogeneity in allelic effect sizes between tumor anatomic sublocations

To characterize tumor subsite-specificity and effect size heterogeneity across tumor subsites for newly identified loci, as well as for established loci associated with overall CRC risk in previous GWAS meta-analyses, we examined the association evidence in three different ways. First, for each index variant we created forest plots of OR estimates with 95% CIs from the GWAS meta-analyses for proximal colon, distal colon, and rectal cancer. Second, we tested for heterogeneity using multinomial logistic regression analysis. In brief, after pooling of data sets, we performed a likelihood ratio test comparing a model in which the ORs for the risk variant were allowed to vary across tumor subsites, to a model in which the ORs were constrained to be the same across tumor sites. Third, we used a multinomial logistic regression-based model selection approach to assess which configuration of tumor subsites is most likely to be associated with a given variant. For each variant, we defined and fitted 11 possible causal risk models specifying variant effect configurations that vary or are constrained to be equal among subsets of tumor subsites (supplementary table 2). We then identified and report the best fitting model using the Bayesian Information Criterion (BIC). For each model *i* we calculated ΔBIC*_i_* = BIC*_i_* − BIC_min_, where BIC_min_ is the BIC value for the best model. Models with ΔBIC*_i_* ≤ 2 were considered to have substantial support and indistinguishable from the best model.[17] For these variants, we do not report a single best model. Analyses were carried out using the VGAM R package.[18] The list of index variants for previously published CRC risk signals is based on Huyghe *et al*.[8].

### Genomic annotation of new GWAS loci and gene prioritization

We annotated all new risk loci with five types of functional and regulatory genomic annotations: (i) cell-type-specific regulatory annotations for histone modifications and open chromatin, (ii) nonsynonymous coding variation, (iii) evidence of transcription factor binding, (iv) predicted functional impact across different databases for non-coding and coding variants, (v) co-localization with eQTL signals. Genes were further prioritized based on biological relevance, colorectal tissue expression, the presence of associated non-synonymous coding variants predicted to be deleterious, evidence from laboratory-based functional studies, somatic alterations, or familial syndromes linking them to CRC or cancer pathogenesis. Detailed methods and references of the databases queried are provided in the online supplementary materials.

## RESULTS

The final analyses included data for 48,214 CRC cases and 64,159 controls of European ancestry. To discover new loci and genetic risk variants with site-specific allelic effects, we conducted five genome-wide association scans of CRC case subgroups defined by the location of their primary tumor within the colorectum: proximal colon cancer (*n*=15,706), distal colon cancer (*n*=14,376), rectal cancer (*n*=16,212), colon cancer, in which we omitted rectal cancer cases, (*n*=32,002), and distal/left-sided CRC, in which we combined distal colon and rectal cancer cases, (*n*=30,588). Next, we systematically characterized heterogeneity of allelic effects between primary tumor subsites for new and previously identified CRC risk variants to identify loci with shared and site-specific allelic effects.

### New colorectal cancer risk loci

Across the five CRC case subgroup GWAS meta-analyses, a total of 11,947,015 SNVs were analyzed. Inspection of genomic control inflation factors (λ_GC_ and λ_1000_) and quantile-quantile (QQ) plots of test statistics indicated no residual population stratification issues (online supplementary materials and supplementary figure 2). Across CRC tumor subsites, we identified 13 CRC risk loci that mapped outside of regions previously implicated by GWASs for overall CRC risk (closest known locus 3.1 megabases away) and that passed the genome-wide significance threshold of *P* <5×10^−8^ in at least one of the meta-analyses (table 1; figure 1; supplementary figures 3 and 4). Seven of these 13 new loci passed a Bonferroni-adjusted genome-wide significance threshold correcting for the five case subgroups analyzed (table 1). All lead variants were well imputed (minimum average imputation *r*^2^ of 0.788), had MAF >1%, and displayed no significant heterogeneity between genotyping sample sets (Cochran’s Q test for heterogeneity *P* >0.05; table 1).

**Table 1.**
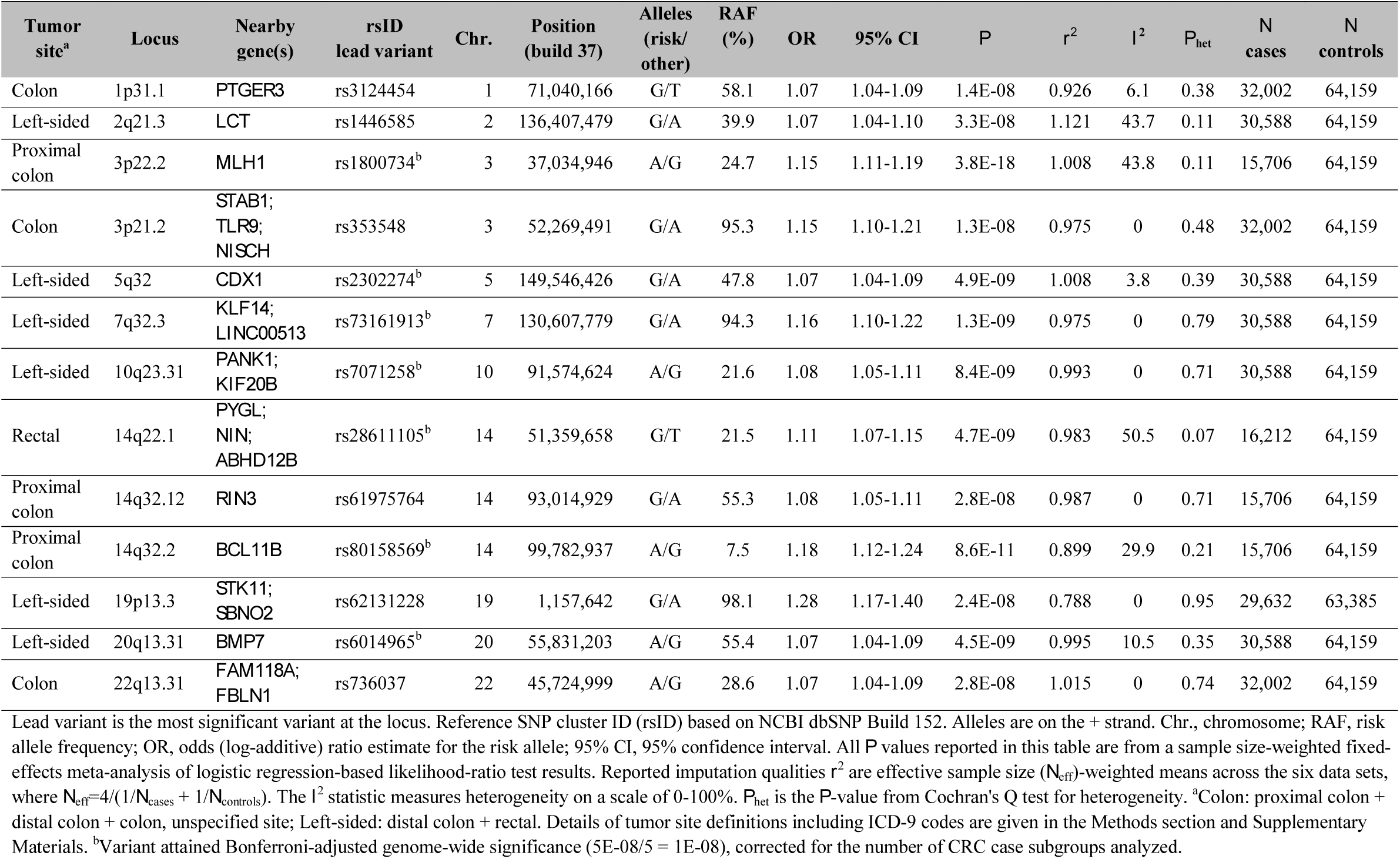
New genome-wide significant colorectal cancer risk loci identified by genome-wide association analysis of case subgroups defined by primary tumor anatomic subsite.

**Figure 1.**
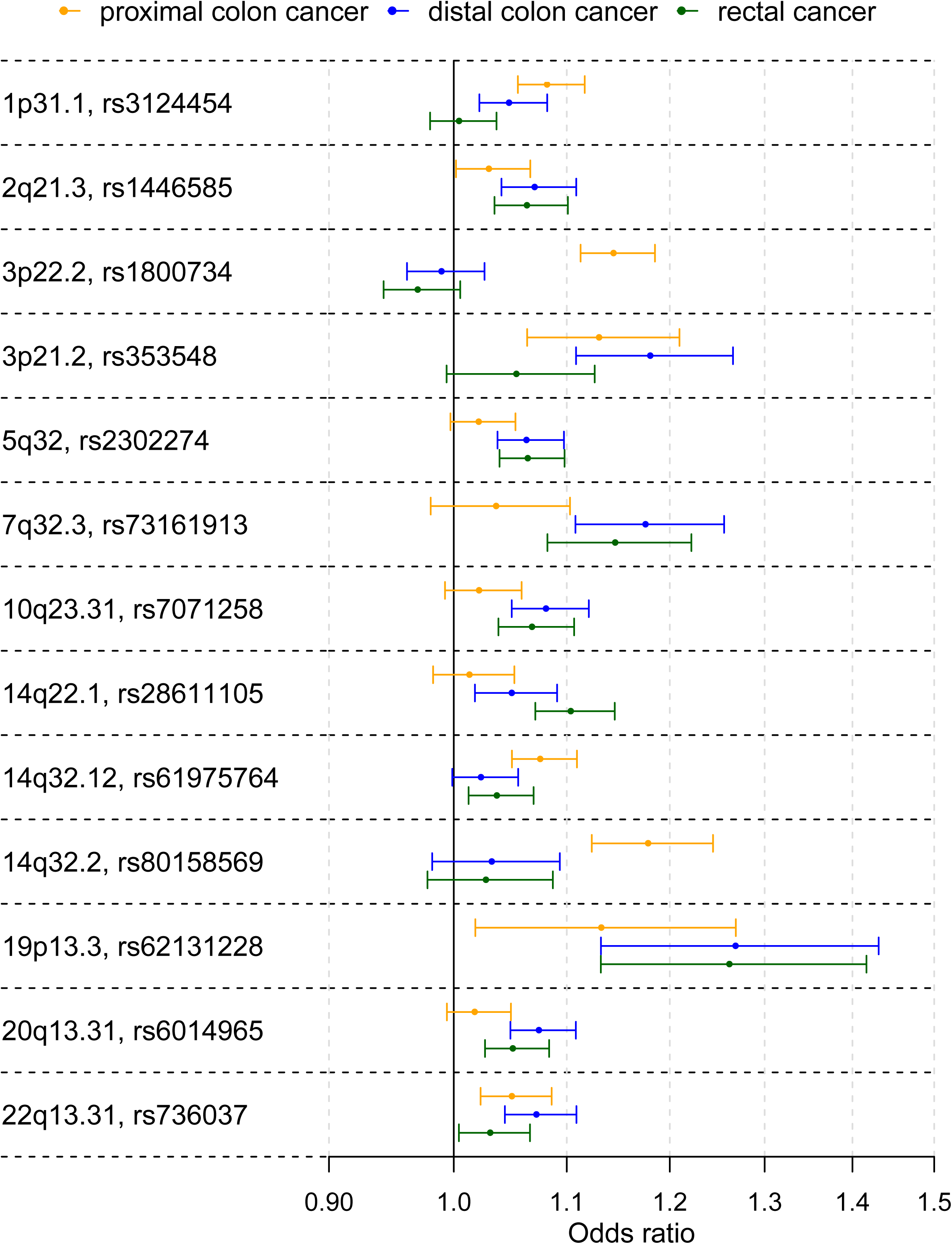
Primary tumor site-specific associations for the lead SNPs of the 13 CRC risk loci not reported in previous GWAS. The forest plots show the (log-additive) odds ratio estimates together with 95% confidence intervals. For clarity, this figure only shows results for the proximal colon, distal colon, and rectal cancer case subgroup analyses.

The novel associations showing the strongest statistical evidence were obtained for proximal colon cancer and mapped near *MLH1* on 3p22.2 (rs1800734, *P*=3.8×10^−18^) and near *BCL11B* on 14q32.2 (rs80158569, *P*=8.6×10^−11^). These loci showed strongly proximal cancer-specific associations. The proximal colon analysis yielded an additional locus at 14q32.12 (rs61975764, *P*=2.8×10^−8^) that showed attenuated effects for cancers at other sites of the colorectum (figure 1 and supplementary table 3). Most new loci (six) were discovered in the left-sided CRC analysis: 2q21.3 (rs1446585, *P*=3.3×10^−8^), near *CDX1* on 5q32 (rs2302274, *P*=4.9×10^−9^), near *KLF14* on 7q32.3 (rs73161913, *P*=1.3×10^−9^), 10q23.31 (rs7071258. *P*=8.4×10^−9^), 19p13.3 (rs62131228, *P*=2.4×10^−8^), and near *BMP7* on 20q13.31 (rs6014965, *P*=4.5×10^−9^). The rectal cancer analysis identified an additional locus near *PYGL* on 14q22.1 (rs28611105, *P*=4.7×10^−9^) that showed an attenuated effect for distal colon cancer (figure 1 and supplementary table 3). No additional new loci were detected in the distal colon analysis. The colon cancer analysis identified three new genome-wide significant loci for colon cancer near *PTGER3* on 1p31.1 (rs3124454, *P*=1.4×10^−8^), 3p21.2 (rs353548, *P*=1.3×10^−8^), and 22q13.31 (rs736037, *P*=2.8×10^−8^).

### Genomic annotations and most likely target gene(s) at new loci

To gain insight into the molecular mechanisms underlying the new association signals, and to identify candidate causal variants and the most likely target gene(s), we annotated signals with functional and regulatory genomic annotations, assessed colocalization with expression quantitative trait loci (eQTLs), and performed literature-based gene prioritization. Notable and strong candidate causal variants and target genes are summarized here. Full results for all new signals are given in supplementary tables 4 and 5.

At the *MLH1* gene promoter region on 3p22.2, associated to proximal colon cancer risk, previous studies have reported strong and robust associations between the common SNP rs1800734, and sporadic CRC cases with high microsatellite instability (MSI-H) status.[19,20] Rare deleterious nonsynonymous germline mutations in the DNA mismatch repair (MMR) gene *MLH1* are a frequent cause of Lynch syndrome (OMIM #609310). The risk allele of the likely causal SNP rs1800734 is strongly associated with *MLH1* promoter hypermethylation and loss of MLH1 protein in CRC tumors.[20] The mechanisms of *MLH1* promoter hypermethylation and subsequent gene silencing may account for most sporadic CRC tumors with defective DNA MMR and MSI-H.[21]

At the highly localized, strongly proximal colon-specific association signal on 14q32.2, the lead SNP rs80158569 is located in a colonic crypt enhancer region and overlaps with multiple transcription factor binding sites, making it a strong candidate causal variant. The nearby gene *BCL11B* encodes a transcription factor that is required for normal T cell development,[22,23] and that has been identified as a SWI/SNF complex subunit.[24] *BCL11B* acts as a haploinsufficient tumor suppressor in T-cell acute lymphoblastic leukemia (T-ALL).[25,26] Experimental work reported by Sakamaki *et al*. suggests that impairment of Bcl11b promotes intestinal tumorigenesis in mice and humans through deregulation of the β-catenin pathway. [27]

At locus 14q32.12, lead SNP rs61975764 showed the strongest evidence of statistical association in the proximal colon analysis and attenuated effects for the other CRC tumor locations. Genotype-Tissue Expression (GTEx) data show that rs61975764 is an eQTL for gene Ras And Rab Interactor 3 *(RIN3)* in transverse colon tissue, the risk allele G being associated with decreased expression. RIN3 functions as a RAB5 and RAB31 guanine nucleotide exchange factor involved in endocytosis.[28,29]

At locus 5q32, one of six loci identified in the left-sided CRC analysis, the intestine-specific transcription factor caudal-type homeobox 1 *(CDX1)* encodes a key regulator of differentiation of enterocytes in the normal intestine and of CRC cells. CDX1 is central to the capacity of colon cells to differentiate and promotes differentiation by repressing the polycomb complex protein BMI1 which promotes stemness and self-renewal. Colonic crypt cells express BMI1 but not CDX1. The repression of BMI1 is mediated by microRNA-215 which acts as a target of CDX1 to promote differentiation and inhibit stemness.[30] Consistent with this view, CDX1 has been shown to inhibit human colon cancer cell proliferation by blocking β-catenin/T-cell factor transcriptional activity.[31]

In a region of extensive LD on locus 2q21.1, lead SNP rs1446585, associated with left-sided CRC, is in strong LD with the functional SNP rs4988235 (LD r^2^ = 0.854) in the cis-regulatory element of the lactase gene. In Europeans, the rs4988235 genotype determines the autosomal dominant lactase persistence phenotype, or the ability to digest the milk sugar lactose in adulthood. The P-value for functional SNP rs4988235 when assuming an additive model was 7.0×10^−7^. The allele determining lactase persistence (T) is associated with a decreased risk of CRC. This is consistent with a previous candidate study that reported a significant association between low lactase activity defined by the CC genotype and CRC risk in the Finnish population.[32] The protective effect conferred by the lactase persistence genotype is likely mediated by dairy products and calcium which are known protective factors for CRC.[33] Of note, the CC genotype has also been associated with a lower body mass index (BMI),[34] presumably because of the nutritional advantage associated with lactase persistence. Since this is a dominant trait with the rs4988235 CC genotype defining lactose intolerance, we also tested left-sided CRC association for these variants assuming a dominant model. Consistent with a dominant model, associations for rs1446585 and rs4988235 became more significant with P-values of 4.4×10^−11^ and 1.4×10^−9^, respectively. For the functional SNP rs4988235, the OR estimate for having genotype CC versus CT or TT, and left-sided CRC was 1.14 (95% CI: 1.09-1.19). Because this region has been under strong selective pressure, it is particularly prone to population stratification and follow-up studies are therefore warranted.[35] However, the fact that we included genotype principal components in the models for all analyzed sample sets, and that the association shows a consistent direction of effect across sample sets (supplementary table 6), suggest that this result is not driven by population stratification.

Candidate genes at the left-sided CRC risk loci 7q32.2 and 20q13.31 are involved in TGF-β signaling. At 7q32.3, the Krüppel-like factor 14 *(KLF14)* gene is a strong candidate. We previously reported loci at known CRC oncogene *KLF5* and at KLF2. [8] The imprinted gene *KLF14* shows monoallelic maternal expression, and is induced by TGF-β to transcriptionally corepress the TGF-beta receptor II *(TGFBR2)* gene.[36] A cis-eQTL for *KLF14*, that is uncorrelated with our lead SNP rs73161913, acts as a master regulator related to multiple metabolic phenotypes,[37,38] and an independent variant in this region has been associated to basal cell carcinoma.[39] For both reported associations, the effects depended on the parent-of-origin of the risk alleles. The association with metabolic phenotypes also depended on sex. We did not find any evidence for the presence of strong sex-dependent effects (males: OR=1.13, 95% CI=1.07-1.20, *P*=4.4×10^−5^; females: OR=1.17, 95% CI=1.09-1.25, *P*=5.4×10^−6^). Further investigation of this locus is warranted to analyze parent-of-origin effects on CRC risk, which is not possible in our dataset. At 20q13.31, the Bone Morphogenetic Protein 7 (BMP7) gene is a strong candidate. In normal intestinal cell crypts, various gradients of TGF-β family members interact with the antagonistic Wnt signaling pathway to maintain homeostasis. Members of the TGF-β family, including several bone morphogenetic proteins (BMPs), frequently have somatic mutations in sporadic CRC tumors, have been implicated by GWASs, and germline mutations are causative for familial CRC syndromes.[40] BMP7 signaling in TGFBR2-deficient stromal cells promotes epithelial carcinogenesis through SMAD4-mediated signaling.[41] In CRC tumors, BMP7 expression correlates with parameters of pathological aggressiveness such as liver metastasis and poor prognosis.[42]

On 14q22.1, the single locus identified only in the rectal cancer analysis, GTEx data show that, in gastrointestinal tissues, colocalizes with a cis-eQTL co-regulating expression of genes *PYGL, ABHD12B*, and *NIN*. Glycogen Phosphorylase L *(PYGL)* is the strongest candidate. We recently identified and replicated an association between genetically predicted *PYGL* expression and CRC risk in a transcriptome-wide association study that used transverse colon tissue transcriptomes and genotypes from GTEx to construct prediction models.[43] Favaro *et al*. showed that this glycogen metabolism gene plays an important role in sustaining proliferation and preventing premature senescence in hypoxic cancer cells.[44] In different cancer cells lines, silencing of *PYGL*, expression of which is induced by exposure to hypoxia, led to increased glycogen accumulation and increased reactive oxygen species levels that contributed to p53-dependent induction of senescence and impaired tumorigenesis.[44]

At new locus 1p31.1, identified in the analysis for colon cancer, *PTGER3* encodes Prostaglandin E Receptor 3, a receptor for prostaglandin E2 (PGE2), a potent pro-inflammatory metabolite that is biosynthesized by Cyclooxygenase-2 (COX-2). COX-2 plays a critical role in mediating inflammatory responses that lead to epithelial malignancies and its expression is induced by NF-κβ and TNF-α. The anti-inflammatory activity of nonsteroidal anti-inflammatory drugs (NSAIDs) such as aspirin and ibuprofen operates mainly through COX-2 inhibition, and long-term NSAID use decreases incidence and mortality from CRC.[45] Prostaglandin E2 (PGE2) is required for the activation of β-catenin by Wnt in stem cells,[46] and promotes colon cancer cell growth.[47] Prostaglandin E Receptor 3 plays an important role in suppression of cell growth and its downregulation was shown to enhance colon carcinogenesis.[48] Hypermethylation may contribute to its downregulation in colon cancer.[48]

### Risk heterogeneity between tumor anatomical sublocations

Multinomial logistic regression modeling of 96 known and 13 newly identified risk variants showed the presence of substantial risk heterogeneity between cancer in the proximal colon, distal colon, and rectum. For 61 variants, the heterogeneity *P*-value (*P*_het_) was not significant (*P*_het_ >0.05). For 51 of those variants, a multinomial model in which ORs were identical for the three cancer sites provided the best fit, and for 8 of the remaining 10 variants, this model did not significantly differ from the best fitting model (supplementary tables 2, 3 and 7; supplementary figure 5).

Among the 109 known or newly discovered variants, 48 showed at least some evidence of heterogeneity with *P*_het_ <0.05, and after Holm-Bonferroni correction for multiple testing, 14 variants showing strong evidence of heterogeneity remained significant (all *P*_het_ <4.6×10^−4^). These included 10 variants previously reported in GWAS for overall CRC risk.

For 17 out of the 48 variants with *P*_het_ <0.05, the best-fitting model supported an effect limited to left-sided CRC (figure 2 and supplementary tables 3 and 7). Of these 17 variants, six were in the list of variants with the strongest evidence of heterogeneity (*P*_het_ <4.55×10^−4^), including the following previously reported loci: *C11orf53-COLCA1-COLCA2* on 11q23.1 (*P*_het_=6.0×10^−14^), *APC* on 5q22.2 (*P*_het_=2.3×10^−10^), *GATA3* on 10p14 (*P*_het_=1.7×10^−8^), *CTNNB1* on 3p22.1 (*P*_het_=9.8×10^−8^), *RAB40B-METRLN* on 17q25.3 (*P*_het_=3.6×10^−6^), and *CDKN1A* on 6p21.2 (*P*_het_=1.6×10^−4^). Inspection of forest plots and association evidence also suggest stronger risk effects for left-sided tumors for the following additional five known loci: *TET2* on 4q24, *VTI1A* on 10q25.2, two independent signals near *POLD3* on 11q13.4, and *BMP4* on 14q22.2.

**Figure 2.**
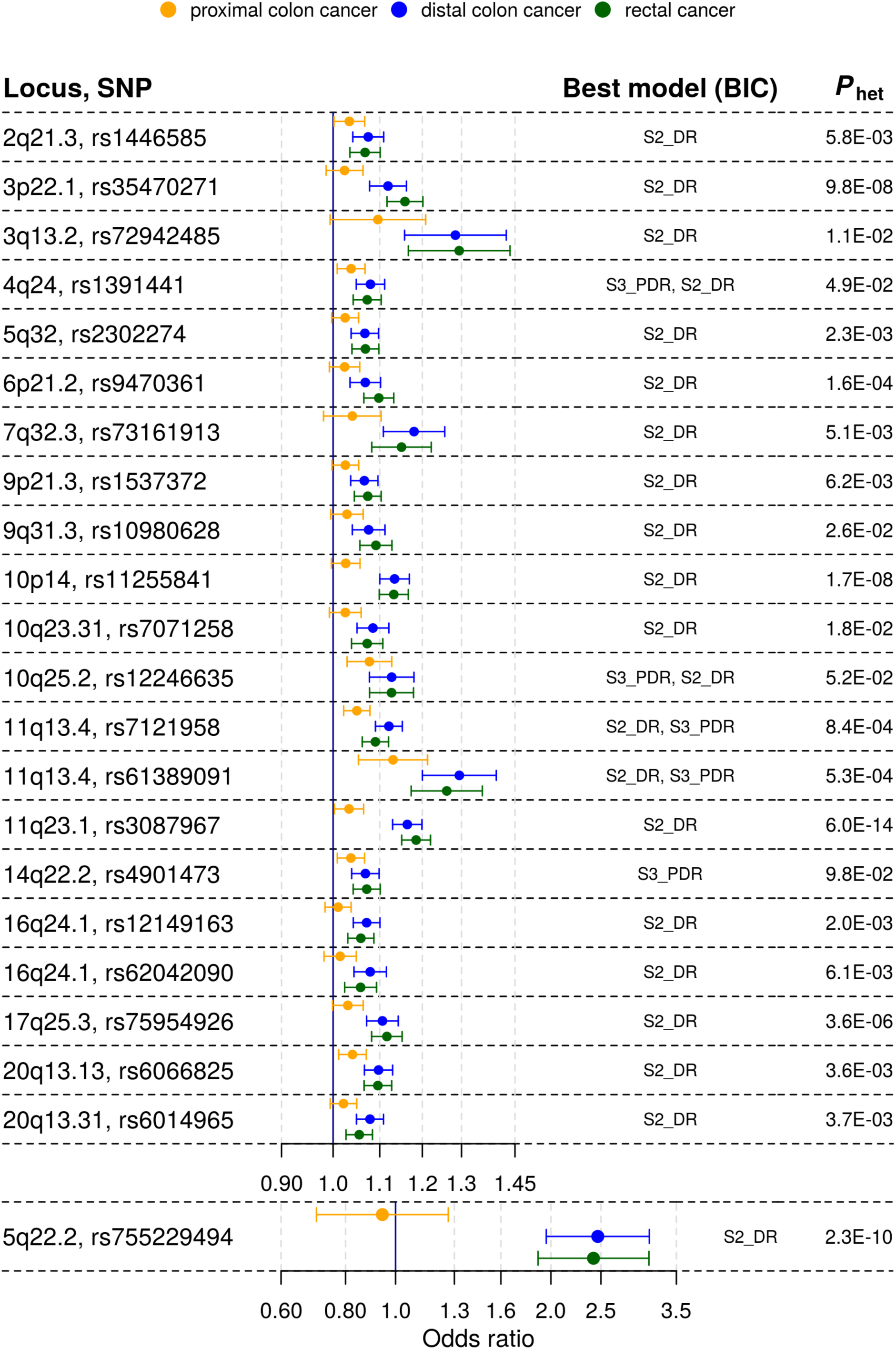
Loci showing association with risk of distal colorectal cancer (i.e., distal colon + rectal), but attenuated or no evidence for association with proximal colon cancer risk. The forest plots show the (logadditive) odds ratio estimates together with 95% confidence intervals from the GWAS meta-analyses of case subgroups defined by primary tumor anatomical subsite for proximal colon, distal colon and rectal. Best model is the best-fitting multinomial logistic regression model according to the Bayesian Information Criterion (BIC). Models are defined in supplementary table 2. *P*_het_ is the *P*-value from a test for heterogeneity of effects across tumor subsites.

For 5 out of the 48 variants with *P*_het_ <0.05, a model with association with colon cancer risk, but no association with rectal cancer risk, provided the best fit (supplementary tables 3 and 7). These involve the following loci: *PTGER3* on 1p31.1, *STAB1-TLR9* on 3p21.2, *HLA-B-MICA/B-NFKBIL1-TNF* on 6p21.33, *NOS1* on 12q24.22, and *LINC00673* on 17q24.3. Association evidence also suggest stronger risk effects for colon tumors for one of two independent signals near *PTPN1* on 20q13.13.

Evidence from the three approaches (figure 1; supplementary tables 3 and 7) indicates that only two loci are strongly proximal colon cancer-specific: *MLH1* on 3p22.2 (*P*_het_=5.4×10^−19^), and *BCL11B* (*P*_het_=1.5×10^−5^) on 14q32.2. Finally, for only 1 variant, at one of two independent loci near *SATB2* on 2q33.1, a model with a rectal cancer-specific association provided the best fit, but association evidence shows attenuated effects for proximal and distal colon cancer. OR estimates also suggest stronger risk effects for rectal cancer at the known loci *LAMC1* on 1q25.3, and *CTNNB1* on 3p22.1, and at new locus *PYGL* on 14q22.1.

## DISCUSSION

It has long been recognized that sporadic CRCs arising in different anatomical segments of the colorectum differ in age- and sex-specific incidence rates, clinical, pathological and tumor molecular features. However, our understanding of the etiological factors underlying these medically important differences has remained scarce. This study aimed to examine whether the contribution of common germline genetic variants to sporadic CRC carcinogenesis differs by anatomical sublocation. The large sample size comprising 112,373 European-ancestry CRC cases and controls provided adequate statistical power to discover new loci and genetic variants with risk effects limited to tumors for certain anatomical subsites, and to compare allelic effect sizes of genetic variants across anatomical subsites.

Our CRC case subgroup meta-analyses identified 13 additional genome-wide significant CRC risk loci that, due to the presence of substantial allelic effect heterogeneity between anatomical subsites, were not detected in larger previously published GWAS for overall CRC risk.[8,9] In fact, the only way to discover certain loci and genetic risk variants with case subgroup-specific allelic effects is via analysis of homogeneous case subgroups.[49] For example, *P*-values for rs1800734 and rs80158569 were ~18 and ~5 powers of ten, respectively, more significant in the proximal colon analysis compared to in our overall CRC analysis. While follow-up laboratory studies are needed to uncover the causal variant(s), the biological mechanism and the target gene, multiple lines of evidence support strong candidate target genes at many of the newly identified genome-wide significant loci, including genes *PTGER3, LCT, MLH1, CDX1, KLF14, PYGL, RIN3, BCL11B*, and *BMP7*.

Previous GWASs had already reported allelic effect heterogeneity between tumor sites, including for 10p14, 11q23, and 18q21 but only contrasted colon and rectal tumors, without distinguishing between proximal and distal colon.[50,51] The sample size and timing of the present study enabled a systematic characterization of heterogeneity of allelic effects between more primary tumor anatomical sublocations, and for a much expanded catalog of CRC risk variants. Our analysis revealed substantial and previously unappreciated allelic effect heterogeneity between proximal and distal CRC. The results further suggest that distal colon and rectal cancer have very similar germline genetic etiologies. Our findings at several loci are consistent with CRC tumor molecular studies. The consensus molecular subtypes (CMSs), which are based on tumor gene expression profiles, are differentially distributed between proximal and distal CRCs. The canonical CMS (CMS2) is highly enriched in distal CRC (56% versus 26% for proximal CRC) and is characterized by strong upregulation of Wnt downstream targets.[52] We found that risk variant associations near Wnt/β-catenin pathway genes *APC* and *CTNNB1* were confined to distal CRC. We also found that associations for variants near genes *BOC* and *FOXL1*, members of the Hedgehog signaling pathway, were confined to distal CRC risk, suggesting that the antagonistic Wnt and Hedgehog signaling pathways may contribute more to the development of distal CRC tumors.

The precise intrinsic or extrinsic effect modifiers explaining the observed allelic effect heterogeneity between anatomical subsites remain unknown and further research is needed. Short-chain fatty acids (SCFAs), in particular butyrate, produced by microbiota through fermentation of dietary fiber in the human colon may be involved. Concentrations of butyrate, which plays a multifaceted antitumorigenic role in maintaining gut homeostasis, are much higher in the proximal colon.[53] Moreover, the known chemopreventive role of butyrate may involve the modulation of signaling pathways including TGF-β and Wnt.[54] This may contribute to possible differences between anatomical subsites in colorectal crypt cellular dynamics with increased stem cell cycling in the distal colorectum promoting growth of precancerous conventional adenomas.

One limitation of our study is that we have not performed discovery GWAS analyses of CRC case subgroups based on more detailed anatomical locations beyond proximal colon, distal colon and rectum. However, given our current total sample size, such analyses would inevitably result in reduced statistical power for new discovery owing to the reduced sample sizes and the aggravated multiple testing burden. As another limitation, our study was based on subjects of European-ancestry and it remains to be determined whether findings are generalizable to other ancestry groups.

In conclusion, germline genetic data support the idea that proximal and distal colorectal cancer have partly distinct etiologies. Our results also demonstrate that future CRC germline genetic studies should take into consideration the differences between primary tumor anatomical subsites. A better understanding of the differing carcinogenic mechanisms and neoplastic transformation risk in the proximal and distal colorectum can inform the development of novel precision treatment and precision prevention strategies through the discovery of novel drug targets and repurposable drug candidates for chemoprevention and cancer treatment, and improved individualized screening recommendations based on risk prediction models that incorporate tumor anatomical subsite.

## Data Availability

All genotype data analyzed in this study have been previously published and have been deposited in the database of Genotypes and Phenotypes (dbGaP), which is hosted by NCBI, under accession numbers phs001415.v1.p1, phs001315.v1.p1, and phs001078.v1.p1. The UK Biobank resource was accessed through application number 8614. CRC-relevant epigenome data were retrieved from the NCBI Gene Expression Omnibus (GEO) database under accession numbers GSE77737 and GSE36401.

## FUNDING AND ACKNOWLEDGEMENTS

Funding statements and acknowledgements are given in the supplemental text.

## Notes

**Disclosures:** The authors disclose no conflicts of interests.

### Competing Interest Statement

The authors have declared no competing interest.

